# Challenges in the Computational Reproducibility of Linear Regression Analyses: An Empirical Study

**DOI:** 10.64898/2026.04.07.26350286

**Authors:** Lee Jones, Adrian Barnett, Gunter Hartel, Dimitrios Vagenas

## Abstract

**Background:** Reproducibility concerns in health research have grown, as many published results fail to be independently reproduced. Achieving computational reproducibility, where others can replicate the same results using the same methods, requires transparent reporting of statistical tests, models, and software use. While data-sharing initiatives have improved accessibility, the actual usability of shared data for reproducing research findings remains underexplored. Addressing this gap is crucial for advancing open science and ensuring that shared data meaningfully support reproducibility and enable collaboration, thereby strengthening evidence-based policy and practice.

**Methods:** A random sample of 95 *PLOS ONE* health research papers from 2019 reporting linear regression was assessed for data-sharing practices and computational reproducibility. Data were accessible for 43 papers. From the randomly selected sample, the first 20 papers with available data were assessed for computational reproducibility. Three regression models per paper were reanalysed.

**Results:** Of the 95 papers, 68 reported having data available, but 25 of these lacked the data required to reproduce the linear regression models. Only eight of 20 papers we analysed were computationally reproducible. A major barrier to reproducing the analyses was the great difficulty in matching the variables described in the paper to those in the data. Papers sometimes failed to be reproduced because the methods were not adequately described, including variable adjustments and data exclusions.

**Conclusion:** More than half (60%) of analysed studies were not computationally reproducible, raising concerns about the credibility of the reported results and highlighting the need for greater transparency and rigour in research reporting. When data are made available, authors should provide a corresponding data dictionary with variable labels that match those used in the paper. Analysis code, model specifications, and any supporting materials detailing the steps required to reproduce the results should be deposited in a publicly accessible repository or included as supplementary files. To increase the reproducibility of statistical results, we propose a Model Location and Specification Table (MLast), which tracks where and what analyses were performed. In conjunction with a data dictionary, MLast enables the mapping of analyses, greatly aiding computational reproducibility.

## Introduction

Concerns about the reproducibility of research findings have attracted considerable attention across multiple scientific disciplines, including psychology, economics, and the health and biomedical sciences, where published results are often difficult to reproduce independently [1, 2, 3]. Empirical assessments of computational reproducibility in health related research have shown that even when original datasets are available, reproducing published statistical results can be challenging, with discrepancies often arising from reporting errors and insufficient documentation of data management and statistical analyses [4, 5].

A widely cited survey of over 1,500 researchers reported that 52% perceived a “reproducibility crisis” in science, 70% had been unable to reproduce another researcher’s findings, and 50% had failed to reproduce at least one of their own studies [6]. Although the survey was based on self-reported experiences from a voluntary sample and did not directly measure reproducibility rates, it played an important role in catalysing broader discussion about research reproducibility. These concerns have contributed to ongoing calls for improvements in study design, statistical practice, and reporting standards [3].

The reproducibility of results strengthens scientific findings by enabling verification and building trust in research. Allowing others to independently confirm that results are not due to coding or data processing errors is key to identifying analytical errors and supporting the accurate interpretation of findings. Reproducibility differs from replication; the latter involves collecting and analysing new data using the original protocol. Computational reproducibility refers to the ability to reproduce the reported results using the original code, the described statistical tests, the data, and the software environments [7]. Achieving this requires well-documented methods, workflows, and a clearly described or accessible software setup, including version-controlled software and dependencies [8, 7].

The volume of information presented in scientific papers has increased substantially over time [9], alongside increasing statistical complexity in medical research, with methods such as multivariable linear regression now widely used [10]. Despite this growing use, statistical errors remain common, often arising from a limited understanding of statistical methods among researchers [11]. Reviews have reported statistical errors or methodological misuse in up to half of assessed articles [12, 13], and p-value inconsistencies that may alter statistical conclusions have been identified in approximately one in eight papers [14]. With over 1.5 million papers indexed annually in PubMed [15], and a large proportion reporting statistical analyses [16], the scale of the literature and its reliance on statistical methods mean that even minor errors can have wide-reaching implications for the reliability of medical research. Addressing reproducibility is crucial for improving healthcare and ensuring that discoveries are trustworthy and impactful.

While many studies have assessed compliance with data-sharing policies [17], few have examined whether shared data are complete, accurate, and sufficiently documented to enable reproducibility. This research moves beyond assessing data availability to evaluate whether shared datasets are usable for reproducing published statistical analyses. Gaps such as incomplete datasets, errors, or inadequate documentation can undermine the goals of open science, leading to wasted effort and resources when research is built upon potentially incorrect findings. Of greater concern is that flawed analyses may be published and ineffective treatments subsequently implemented in practice. This study evaluates the quality and reproducibility of open datasets, identifies common barriers to reuse, and advocates for standardized practices to promote transparency, consistency, and reproducibility in statistical analysis and reporting in health research.

## Materials and Methods

This exploratory, descriptive cross-sectional study does not aim to test specific hypotheses. Using a random sample of published articles, the study estimates the prevalence of research papers with open data that are computationally reproducible. This work is part of a broader project on statistical reporting and reproducibility in health research, with full details of the paper sampling methods provided in Jones et al. [18].

### Sample size determination

The original sample size of 100 papers was determined to assess the prevalence of reported linear regression assumptions, allowing for a detectable sample proportion of 0.05 (5%) with a two-sided 95% confidence interval and a 5% margin of error [18]. Based on pilot work conducted in 2018, approximately 50% of papers were expected to have associated data. This percentage was considered feasible for assessing computational reproducibility.

### Research questions

- What percentage of papers stating that data were available included the data necessary to reproduce the linear regression analyses?
- Among papers that shared data, what percentage included a data dictionary?
- What percentage of linear regression analyses in published papers were computationally reproducible?

### Randomisation of papers

One hundred research papers (excluding editorials and other non-research articles) were randomly selected from *PLOS ONE* in 2019, a journal with an established data-sharing policy [19]. This study was conducted in 2020/21, with 2019 chosen to provide a current perspective on reporting practices. An unintended benefit of this time frame was that it covered the last full year before the COVID-19 pandemic, thereby avoiding potential disruptions to research practices caused by pandemic-related shutdowns [20]. The “searchplos” function from the “rplos” R package [21] was used to identify papers based on two criteria: “health” appearing in the subject area and “linear regression” reported in the materials and methods section. The eligible papers were randomly ordered, and the first 100 that met the criteria were included. Each paper was identified by a unique study number (paper 1 to 100); the corresponding paper DOIs are provided in GitHub [22]. To maintain a focus on the routine use of standard linear regression among health researchers, studies were excluded if their analyses incorporated clustered or random effects, alternative methods (e.g., Bayesian or non-parametric approaches), or used linear regression for secondary purposes like calibration or pre-processing, rather than as part of their main analysis. Five papers from the original 100 that met the inclusion criteria were excluded upon closer inspection for failing to report any linear regression results, leaving 95 papers for assessment of reporting [18].

Computational reproducibility was assessed for the first 20 papers in the random sample with confirmed data, ensuring the representativeness of the overall study sample. However, we acknowledge that the final sample size was small. Reproducing each paper was time-intensive, requiring between one and three days per paper. All reproductions were undertaken by the first author, an accredited senior biostatistician with 20 years’ experience, with input, where needed, from three senior biostatisticians (co-authors), two of whom were also accredited (AStat).

### Ethics and Protocol

The aim of this study was to reproduce the statistical results from publications that made their data publicly available. Authors of papers published in *PLOS ONE* were considered to have consented through their agreement with the journal’s data sharing policy [19], which supports the validation and reproduction of results from shared data. This study received Negligible-Low Risk Ethics approval from the Queensland University of Technology Human Research Ethics Committee (Approval Number: 2000000458). The study protocol is publicly available on GitHub [23].

### Linear regression models within papers

Each linear regression model was identified and assigned a number within each paper. A maximum of three linear regression analyses were selected per paper. The final model (or the primary models of interest) was selected for papers with more than three models, with additional models chosen at random. If no primary models of interest were identified, all three models were selected randomly. Limiting the selection to three models per paper made the workload manageable. We also expected a high correlation in computational reproducibility across models from the same paper, leading to diminishing returns from reproducing every model.

Identification of linear regression models was guided by a prior statistical review of the same papers, which was based solely on reported results [18]. Linear regressions were recorded for each paper, and two statisticians independently verified the number of eligible models. Poor reporting practices, including unclear methods, incomplete results, and missing test details, made identification challenging [24]. The initial model counts and locations were refined following data review and finalised prior to model selection, with random sampling applied only when no primary model of interest was identified.

### Reproducibility methodology

For reproducibility purposes, a Quarto report was developed for each of the 95 papers to assess data sharing and reproducibility. Each report documented the statistical software used, whether the data were reported, where they were stored, whether the reported data allowed reproduction of the linear regression models, and whether the results could be computationally reproduced. Our reporting for papers, including reproducible code, HTML reports for each paper, and the code and data required to reproduce the current study, are all available on GitHub [22].

When data were available, the identified linear regression models were re-analysed to assess reproducibility [18]. To improve readability and navigation, the report was broken into four sections: *Original Results*, *Reproduced Results*, *Differences*, and *Sensitivity Analysis*. This layout helped structure the findings and allowed users to move efficiently between different aspects of the analysis. The papers’ results were presented as text and tables and entered into the “*Original Results*” section in a structured format, with results left blank if not reported. The same format was used for the reproduced results to facilitate comparisons, and the differences between the original and reproduced results were graphed and tabulated.

Models were reproduced using the *linReg* function from the JMV package [25] in R. This package was chosen for two reasons: it provides well-formatted output equivalent to the results of other major packages such as SPSS and STATA, and it was also developed for Jamovi [25], a point-and-click version of R that reduces the barrier to reproducibility by allowing users to capture code for their analyses. Differences in coefficients and sums of squares can partly arise from model parameterisation. Reporting was matched to the original study where possible; when sums of squares were not reported, we calculated Type III sums of squares using effect (grand mean) coding [26] to ensure comparability with commonly reported output in major statistical packages [27].

Percentage change was initially considered to assess discrepancies in computational reproducibility; however, it was ruled out early in the process because parameter values near zero produced disproportionately large percentage differences. Transformations of percentage differences were also rejected for their artificial and unintuitive nature. In linear regression, models are expected to produce identical results when reproduced correctly, with any minor discrepancies typically arising from rounding.

A custom function was developed to assess agreement between original and reproduced values for regression coefficients, standard errors, 95% confidence intervals, t-statistics, p-values, R2, adjusted R2, model and residual degrees of freedom, and the global F statistic. For each value, the function computed two thresholds based on the number of decimal places (d) reported in the original paper: a stricter threshold 0.50 *×* 10*−d* and a more lenient threshold (0.999 *×* 10*−d*). Values were classified as “Reproduced” if the absolute difference fell within the stricter threshold, “Incorrect Rounding” if within the lenient threshold, and “Not Reproduced” if exceeding both. This approach provided a structured framework for evaluating reproducibility while accounting for plausible rounding variability.

An example of using these thresholds to evaluate lower confidence intervals is shown in Table 1. In this case, results were reported to two decimal places; therefore, the threshold for considering the lower confidence interval as reproduced was an absolute difference of *≤* 0.005. Absolute differences between *>* 0.005 and 0.0099 were classified as rounding discrepancies. Differences of *≥* 0.01 (in either direction) were flagged as not reproduced. In this example, the lower confidence interval for the intervention group in the original results was reported as 3.77, whereas the reproduced results were *−*3.77664, indicating the original result lacked a negative sign and was therefore not reproduced.

**Table 1.** Example of reproducing lower confidence intervals from a linear regression analysis.

| Term | O_lower | R_lower | Change.lci | Reproduce.lower |
| --- | --- | --- | --- | --- |
| Intercept | 83.52 | 83.5163 | -0.0037 | Reproduced |
| Age | 1.46 | 1.4508 | -0.0092 | Incorrect Rounding |
| BMI: |  |  |  |  |
| Overweight – Normal | 4.71 | 4.7132 | 0.0032 | Reproduced |
| Obese – Normal | 9.5 | 9.4956 | -0.0044 | Reproduced |
| Group: |  |  |  |  |
| Intervention – Control | 3.77 | -3.7664 | -7.5364 | Not Reproduced |

O\_lower = original lower confidence interval; R\_lower = reproduced lower confidence interval; change.lci = change in R\_lower - O\_lower; Reproduce.lower = lower confidence interval reproduced.

The study protocol [23] guided the development of the reproducibility template. ChatGPT [28] was used to aid in developing functions and debugging code, while both Grammarly [29] and ChatGPT were used to edit grammar and improve writing flow. However, neither tool was used to write sections of the manuscript or perform statistical reviews.

Data were imported directly from the *PLOS ONE* site or associated data repositories using code that extracted the necessary information without storing entire datasets. The relevant regression models were identified and reformatted for each paper into a standardized structure, dynamically renaming predictors, e.g., *age* becomes *x1* in the formula, but is still labelled as *age* in the output. Modular functions were created to reproduce the analyses and assess the difference between the original and reproduced results.

Up to three linear regression models were assessed per paper. At the model level, computational reproducibility was categorised into four levels. Model-level classifications were based on expert assessment of the extent and nature of the discrepancies identified, rather than on a predefined percentage of model terms reproduced, with particular consideration given to whether discrepancies may affect the conclusions of the original analysis:

- **Reproduced**; all aspects of the model were reproduced without errors.
- **Mostly reproduced**; most aspects of the models were reproduced, with errors that did not change the conclusion.
- **Partially reproduced**; only some aspects of the model were reproduced, or errors and differences may affect the conclusion.
- **Not reproduced**; all or most aspects of the model could not be reproduced.

A paper was considered reproducible if all three models were evaluated as either fully or mostly reproduced; otherwise, it was classified as not reproduced.

### Statistical methods

Three aspects were assessed for each paper: (i) whether the reported data were available, (ii) whether any data dictionary was provided, and (iii) whether the paper was computationally reproducible. Each aspect was coded as a binary variable (“Yes” or “No”). Frequencies and percentages were reported for these categorical variables, along with 95% Wilson confidence intervals [30]. Reproducibility was further examined at both the individual statistic level and the model level. At the individual statistic level, reproducibility was classified as “Reproduced”, “Incorrect Rounding”, or “Not Reproduced” and visualised using swimmer plots [31].

While probabilities of reproducibility status could be calculated directly from individual statistics, a modelling approach was required to account for the hierarchical structure of the data, with multiple statistics per model and up to three models per paper. Bayesian mixed-effects multinomial models [fitted using the *brms* package 32] with weakly informative priors to stabilise the variance were used to classify models into reproducibility categories [33]. The models included a random intercept for paper to account for the correlation arising from repeated measures. Predicted probabilities and corresponding 95% credible intervals were estimated for classification at both the individual-statistic and model levels.

The primary author also rated the papers’ subjective difficulty to reproduce on a five-point scale: 0 = no models in the paper were at least “Partially reproduced”, 1 = very difficult, 2 = difficult, 3 = manageable, and 4 = straightforward. This rating, along with other categorical variables (e.g., statistical package used, data format, typographic errors, reporting clarity), were summarised using frequencies and percentages. R version 4.4.2 was used for all analyses [34].

## Results

The majority of papers (84%, 80/95) had an observational study design, and most (77%, 73/95) involved human participants (Table 2). All papers were first screened for data availability, the presence of a data dictionary, data format, and the statistical package used. Of the papers screened, 68 reported that data were available in the paper, supporting information, or in an external data repository (Fig 1). However, the data required to reproduce the linear regression could not be identified for 25/68 of these papers. From the papers with available data (N = 43), a subsample of 20 was sequentially assessed for computational reproducibility.

**Figure 1.**
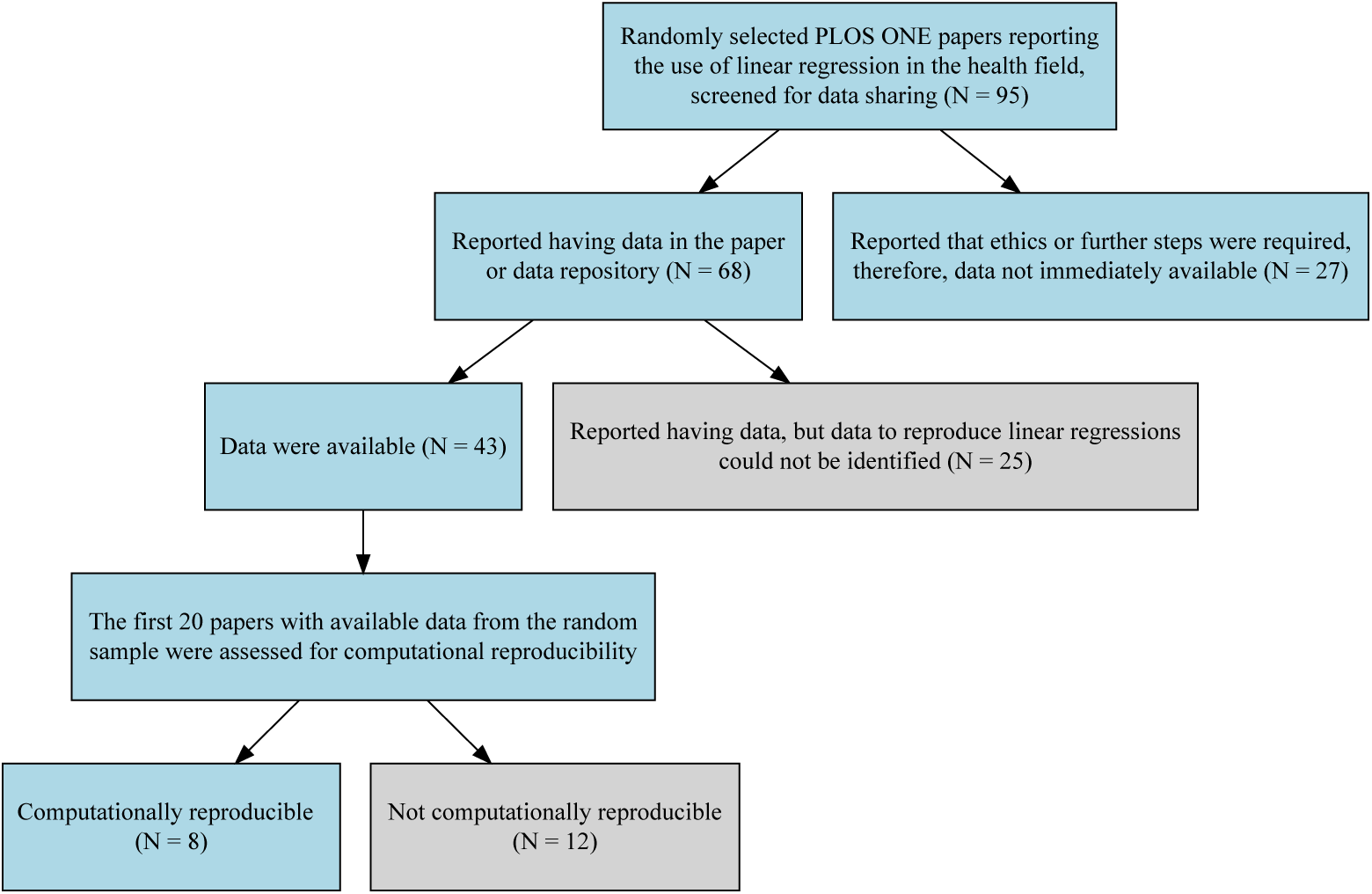
Flow chart of papers in analysis. N is the number of papers.

**Table 2.** Study design and sample type of the 95 included linear regression papers.

| Characteristic | n (%) |
| --- | --- |
| Study Design |  |
| Observational | 80 (84%) |
| Experimental | 15 (16%) |
| Participant Types |  |
| Human | 73 (77%) |
| Animal | 12 (13%) |
| Mix of animal and human | 3 (3%) |
| Mix of animal and plant | 2 (2%) |
| Other studies | 3 (3%) |
| Lab study with comparison to other studies | 1 (1%) |
| Environmental samples | 1 (1%) |

Adapted from Jones et al. p17. [18].

Only 20/43 papers or 47% (95% CI: 33%, 61%) with available data included data dictionaries (Table 3). The most common format for sharing data was Excel (n =29), with a further two authors sharing data using CSV files.

**Table 3.** Frequencies of statistical packages and data formats.

| Statistical package, N = 95 | n (%) | Data Format, N = 43 | n (%) |
| --- | --- | --- | --- |
| SPSS | 33 (35) | Excel | 26 (60) |
| R | 18 (19) | SPSS | 6 (14) |
| Stata | 15 (16) | Stata | 3 (7) |
| Not Reported | 9 (9) | CSV | 2 (5) |
| SAS | 8 (8) | Image | 2 (5) |
| Graphpad Prism | 3 (3) | Website | 2 (5) |
| JMP | 2 (2) | PDF | 1 (2) |
| MATLAB | 2 (2) | Word | 1 (2) |
| PLINK | 2 (2) |  |  |
| BMDP | 1 (1) |  |  |
| Excel | 1 (1) |  |  |
| Statistica | 1 (1) |  |  |

By far the most common statistical package was SPSS, followed by R, Stata and SAS, with less commonly used packages including GraphPad/Prism, MATLAB, and Statistica. Nine authors shared data using statistical programs (Stata = 3, SPSS = 6), these tended to have better structure due to having in-built data dictionaries and in the format required for analysis. A further five papers were in less accessible formats, including PDF, Word or image files and required specialised R packages to extract the data.

### Computational reproducibility results

Of the 20 papers assessed for computational reproducibility, 10 had unclear reporting of the number of observations which complicated the assessment of reproducibility. This was particularly problematic for multivariable models, where it was difficult to determine whether differences were due to uncertainty about which variables were included in the model or to differences in the number of observations. Five papers contained typographical errors such as a missing negative sign, or swapping the order of digits.

At least one model was unclear in nine papers, making the results difficult or not possible to reproduce based on the available information. Overall, eight or 40% (95% CI: 22%, 61%) of papers were found to be computationally reproducible. Only two were rated as straightforward to reproduce, five were rated as difficult, a further three were very difficult to reproduce and six papers had no models that could be at least partially reproduced.

For each paper, we tried to reproduce three regression models. For each regression, we attempted to reproduce multiple terms, including the coefficients (B), CIs, p-values, and test statistics. Similar patterns were typically observed within the same paper. As if a few coefficients could not be reproduced, the others in that paper also failed to reproduce. Conversely, when most coefficients were successfully reproduced but one or two were not, this indicated isolated reporting or typographical errors rather than systematic analytical differences.

Overall, reproduction of individual model statistics was low (Table 4). Only around half of regression coefficients (61/118, 52%) and p-values (72/133, 54%) were reproduced, with lower reproduction for R2 (9/19, 47%), standard errors (6/16, 38%), standardized coefficients (8/22, 36%), and adjusted R2 (3/9, 33%). These observed proportions were influenced by paper-level clustering, as individual papers contributed multiple model statistics and failures to reproduce could occur across several statistics within the same paper. Paper-level clustering was therefore accounted for using multinomial mixed-effects models; the estimated probabilities and 95% credible intervals are reported in Table S1. At the model level, of the 51 models assessed, 12 (24%) were classified as Reproduced, 15 (29%) as Mostly Reproduced, 5 (10%) as Partially Reproduced, and 19 (37%) as Not Reproduced (S2).

**Table 4.** Reproducibility of reported model statistics.

| Terms | Papers | N | Reproduced | Incorrect Rounding | Not Reproduced |
| --- | --- | --- | --- | --- | --- |
|  |  |  | n (%) | n (%) | n (%) |
| B | 16 | 118 | 61 (52) | 7 (6) | 50 (42) |
| Lower CI | 10 | 76 | 46 (61) | 7 (9) | 23 (30) |
| Upper CI | 10 | 76 | 44 (58) | 5 (7) | 27 (36) |
| SE | 2 | 16 | 6 (38) | 4 (25) | 6 (38) |
| t | 0 | 0 | 0 (0) | 0 (0) | 0 (0) |
| p-value | 18 | 133 | 72 (54) | 5 (4) | 56 (42) |
| $R^2$ | 7 | 19 | 9 (47) | 2 (11) | 8 (42) |
| Adjusted $R^2$ | 3 | 9 | 3 (33) | 0 (0) | 6 (67) |
| F | 3 | 9 | 7 (78) | 0 (0) | 2 (22) |
| Standardized B | 2 | 22 | 8 (36) | 0 (0) | 14 (64) |

Terms = model statistics; Papers = number of papers reporting each term; N = total number of times terms were reported across the selected models (up to three per paper).

The reproducibility status of each regression coefficient within each model is displayed by paper in the swimmer plot (Fig 2). For example, in the seventh paper, all 20 regression coefficients across three models were classified as Not Reproduced, with none classified as Reproduced or Incorrect Rounding.

**Figure 2.**
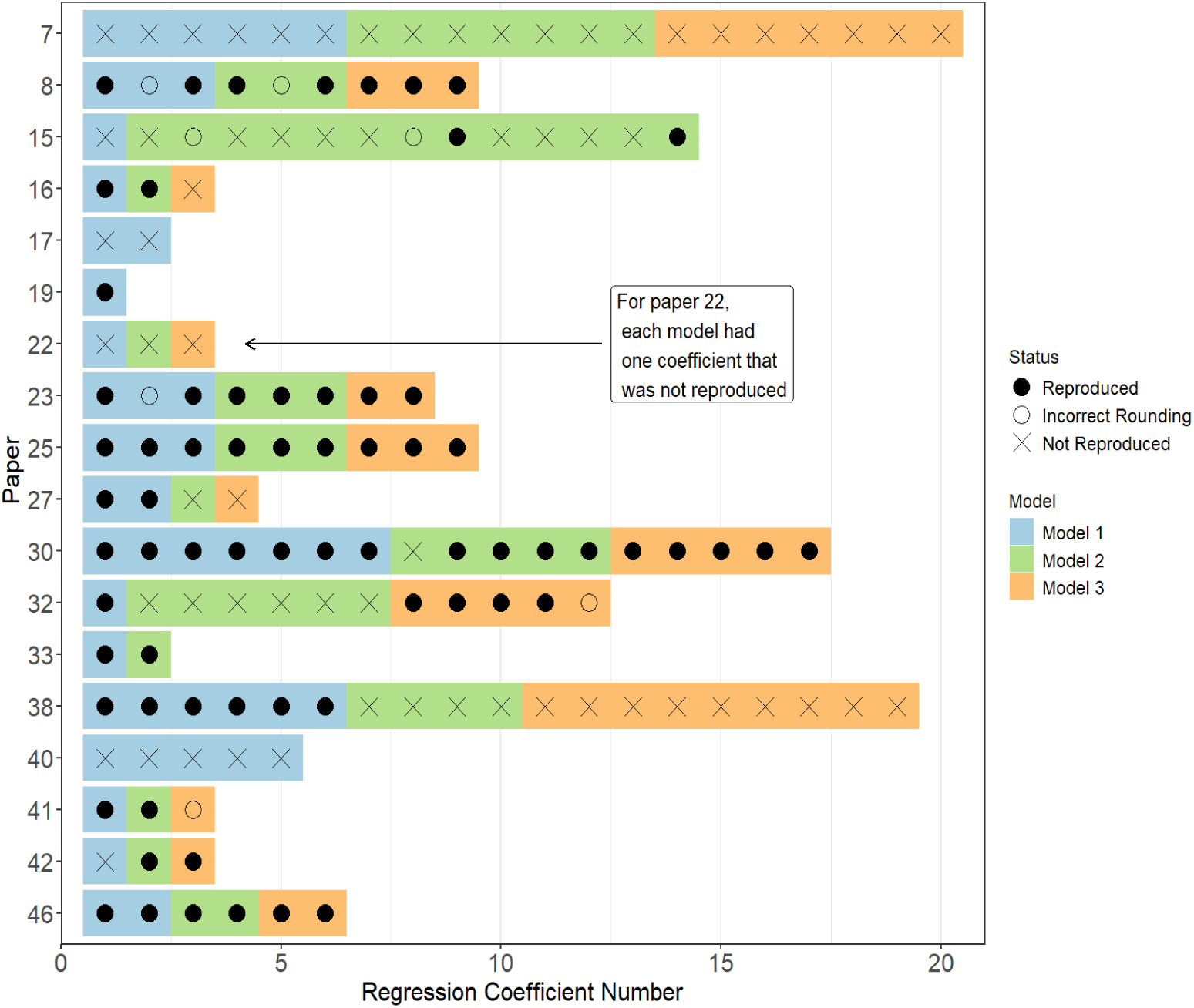
Swimmer plot showing reproducibility status for linear regression coefficients. Black filled circles indicate coefficients that were Reproduced, hollow circles indicate Incorrect Rounding, and crosses indicate Not Reproduced. Blue tiles represent coefficients from the first model, green from the second, and orange from the third model.

The swimmer plot includes 16 papers reporting unstandardized regression coefficients and two papers reporting standardized regression coefficients. Two of the 20 papers assessed are not displayed because regression coefficients could not be evaluated: Paper 44 reported only p-values, which could not be reproduced, while for Paper 12 the available raw data could not be used to reconstruct the data required to fit the reported linear regression model. Both papers were therefore classified as Not Reproduced.

## Discussion

Our findings raise serious concerns about the computational reproducibility of published health research. More than half of the papers assessed failed to be reproduced. These findings align with prior evidence of limited computational reproducibility and reinforce the importance of transparent, detailed reporting of statistical analyses [5, 35]. Understanding and reproducing published analytical methods is important for accurately interpreting results and identifying potentially flawed or incorrect findings before they influence clinical practice, policy, and future research, with potential downstream consequences for patient care and health outcomes.

A key barrier to computational reproducibility was incomplete or unclear methodological reporting, even when data were made available [5, 36]. Problems were often evident in the manuscript before examining the underlying data [24]. In our sample, fewer than half of the papers provided a data dictionary, limiting the ability to identify model variables, interpret coding schemes, and reconstruct derived measures. When key variables or analytic decisions cannot be clearly identified, computational reproduction becomes substantially more challenging and, in some cases, impossible. These findings highlight the need to strengthen researcher training, reporting standards, and review practices [35]. Authors should clearly specify the statistical test conducted, the model used (including parameterisation and assumptions), the software package and version, and any custom functions or code. Full documentation of the dataset, including a data dictionary and all data-processing steps, is essential for computational reproducibility.

One of the difficulties faced in reproducibility research is the lack of an agreed-upon definition of what constitutes reproducibility [37]. One common criticism of computational reproducibility in the literature is the overly rigid definition of exactness, which often fails to account for practical realities such as rounding errors and minor numerical differences arising from using different software (e.g., R vs SPSS) [38]. The appropriate threshold may also depend on the study’s purpose. For example, Seibold et al. [39] examined the reproducibility of longitudinal research focusing on Generalized Estimating Equations (GEE), where exact numerical agreement was not the primary goal, as different software packages often implement GEE differently, making small differences expected and unimportant.

In our study, minor errors were frequently identified in papers for which reproduction was possible, including rounding discrepancies and typographic mistakes. Although these discrepancies typically did not alter the main conclusions, they highlight vulnerabilities in the publication process. Some may reflect systemic pressures faced by researchers, such as time constraints and the pervasive “publish or perish” culture in academia [40]. However, certain errors, for example, point estimates reported outside their corresponding confidence intervals, should have been detectable during peer review or editorial assessment. While many minor discrepancies likely arise during manuscript preparation, our study did not assess the stage at which errors were introduced, leaving open the possibility that some discrepancies may have emerged during journal production. Copy-editing and typesetting involve additional editorial modification, and although such processes are unlikely to account for most errors, variability in editing quality has been noted in academic commentary [41].

Our study also reinforces that while data sharing is a critical step toward transparency, availability alone does not guarantee reproducibility. A substantial proportion of shared datasets lacked sufficient documentation or completeness to support replication of the original analyses. Although some journals, funders, and institutions now mandate or encourage data sharing, these policies have often been evaluated primarily in terms of compliance rates, with less attention to the usability or quality of the shared data [4]. Without clear documentation, accessible data formats, and access to associated code, even datasets labelled as “available” may not enable meaningful verification or further research.

Differentiating levels of reproducibility helped clarify the likely source of observed differences. In some cases, these were attributable to incorrect or missing data, such as differences in reported sample sizes between the manuscript and the available dataset, or to models that did not align with the descriptions provided in the paper. Another plausible explanation for partial reproducibility is that datasets may change throughout the research process. Although we did not assess data management practices directly, datasets often undergo multiple rounds of cleaning, transformation, or filtering, and it is possible that published tables reflected earlier dataset versions rather than the final dataset shared. In such situations, statistics may not align if outputs were not consistently updated following data revisions. Although the exact cause could not always be determined, likely explanations could sometimes be inferred based on the available information. These considerations highlight the importance of maintaining consistent documentation and version control throughout the analysis process.

Several guidelines have been developed to promote reproducibility and transparency, including the FAIR principles (Findable, Accessible, Interoperable, and Reusable) [42], the TOP Guidelines (Transparency and Openness Promotion) [43], and initiatives such as Registered Reports, which emphasise pre-specifying hypotheses and methods to reduce bias [44]. While the development of such frameworks remains important, our findings suggest that the priority should now shift toward strengthening the implementation of these existing principles, particularly the expectations around data sharing and documentation quality.

Improving adherence to open science practices will require coordinated efforts across the research ecosystem [24]. In addition to journal policies, universities and research institutes must actively support reproducibility practices, and funding bodies should continue to embed expectations for data management and sharing into funding requirements. Encouragingly, there is growing international momentum: for example, the US National Institutes of Health (NIH) [45] and the Australian National Health and Medical Research Council (NHMRC) [46] have recently introduced mandatory data sharing plans. Strengthening and evaluating these efforts are critical to achieving transparency and reproducibility in practice, rather than remaining an aspirational goal.

Computational reproducibility represents only one dimension of reproducibility. Statistical results may be numerically reproducible yet still lack scientific or statistical validity [37, 47]. A more complete assessment of scientific reliability also considers inferential reproducibility, that is, whether the modelling approach and its assumptions support the conclusions drawn. In related work [48], we examine inferential reproducibility in the context of linear regression. Attention to both computational accuracy and inferential validity is therefore necessary to ensure that reported findings are sufficiently accurate and precise to guide evidence-based practice.

### Data sharing and transparency

Insufficient or inaccurate reporting of study design and data analysis makes replication challenging, hindering the reliability and cumulative nature of research findings [49, 50]. Poorly reported statistical analyses reduce the reproducibility of individual studies and undermine the scientific effort to build evidence-based practices. With only 40% of papers found to be computationally reproducible, addressing these issues requires systemic changes, such as promoting transparent and rigorous reporting standards, encouraging collaboration with statisticians [51], and emphasising reproducibility as a key component of research quality.

Despite the introduction of data-sharing policies by some journals, the uptake of data sharing has remained limited [17, 52]. A 2023 systematic review examined data and code sharing practices across the medical and health sciences, synthesising findings from 105 meta-research studies covering over 2.1 million articles published between 2016 and 2021 [52]. While 8% of articles declared their data to be publicly available, only 2% were confirmed to be publicly available. Factors associated with greater data sharing included journal policies mandating availability and funding requirements. The authors recommended that policymakers periodically audit publications for compliance [52]. These findings were consistent with our study, which also identified a substantial gap between data availability declarations and the usable, complete raw data provided by the authors.

*PLOS ONE* was selected for the current study because it has promoted open science and data sharing, with policies dating back to 2014 [19]. *PLOS ONE* requires that all data underlying a study’s findings be made fully available at the time of publication, without restriction, except where legal or ethical constraints apply [53]. However, several studies have found that data availability statements are often inconsistent and ambiguous. Ellis et al. [54] argue that vague statements such as “data are available upon reasonable request” may function as a diversion, offering no assurance that data will be provided and lacking a clear definition of what constitutes a “reasonable” request. Poor adherence to data sharing has been documented, for example, Gabelica et al. [55] assessed 1,792 manuscripts published in January 2019 across 333 open-access BioMed Central journals that included a willingness to share data, but found that only 122 (6.8%) actually provided the data upon request.

*PLOS ONE* requires authors to include a Data Availability Statement specifying whether data are available, and if so, where and how they can be accessed (e.g., via a public repository or as supporting information) [19]. An assessment of papers published between the policy’s introduction in March 2014 and May 2016 [56] found that only 20% deposited data in a public repository, the preferred method of the journal. Most authors instead reported that data were available within the paper or its supporting information. However, the study did not assess whether these forms of sharing met the requirements set by the journal’s policy.

Our study identified a large discrepancy between authors’ statements that all data were included in the manuscript or in the supporting information and the actual availability of the raw data. Specifically, 25 out of 68 studies did not provide raw data to reproduce the linear regression analyses. Many of these authors by default reported “All relevant data are within the manuscript and its Supporting Information files”, but in most cases, papers only included analyses tables with summary data. In some cases, no supporting information files were actually available. Some of these discrepancies could have been easily identified through simple automated checks at the submission stage, such as checking for data files.

Cross-disciplinary reviews have noted substantial variation in reproducibility practices, with economics journals typically implementing stronger data and code requirements than health and biomedical journals [3, 57]. Experience from journals of the American Economic Association shows that pre-publication data and code verification can be implemented in routine editorial workflows, although such practices are still rare in health and biomedical research [52, 58].

As part of broader efforts to promote open science and data transparency, *PLOS ONE* implemented several initiatives to improve data sharing and discovery. In 2021, it received a Wellcome Trust grant to support this work [59]. The initiative, first piloted in *PLOS Pathogens*, aimed to integrate data repositories such as Dryad and introduce visual badges to highlight accessible datasets, aligning with FAIR principles (Findable, Accessible, Interoperable, and Reusable) [42]. In 2022, *PLOS* launched the Accessible Data icon, which is automatically applied to articles that link to recognised public repositories and retrospectively applied for papers published since 2016 [59]. However, because the icon relies on detecting repository links in data availability statements, it is only applicable to papers that use external repositories. It does not apply to cases where data are provided via supplementary files. While data badges may be useful for indicating when data are available, they do not address the core issue of data unavailability in the paper or the supporting information, despite the author’s claims that they were provided.

While six papers in our sample had a *PLOS* data badge, one paper correctly stated that individual-level data were not available and provided simulated data. Two additional papers were assessed as not providing the data required to reproduce the linear regression analyses, either because key variables needed to be derived without accompanying description or code, or because the necessary variables for the model were not supplied. Therefore, the data-sharing badge in our sample did not accurately reflect whether the data were sufficient to enable computational reproducibility.

Data badges have been investigated to assess whether their presence increases the likelihood that authors will share their data and code. While the evidence is mixed, some studies suggest that offering data badges as a reward has a limited impact [60, 61]. In a randomised controlled trial involving 80 author teams per group, Rowhani-Farid et al. [61] found that offering a data sharing badge did not significantly increase data sharing, with similar odds of sharing between intervention and control groups (odds ratio = 0.9, 95% CI: 0.1, 9.0); only two teams in each group provided data.

In a separate study evaluating the outcomes of a badge policy rather than its causal effect, Crüwell et al. [60] assessed the computational reproducibility of 14 published articles. Although all articles provided at least some data and six provided analysis code, only one was exactly reproducible and three were essentially reproducible with minor deviations. Together, these studies suggest that incentive-based mechanisms like badges may be insufficient on their own. Crüwell et al. [60] recommended that journals implement reproducibility checks during peer review rather than relying solely on author disclosure. This aligns with our study, which found that authors’ disclosure of shared data is a poor substitute for validation on submission.

### Practical recommendations

Verifying that data are available when claimed in the methods section should be a standard part of compliance checks. When data are shared, a data dictionary must also be required to facilitate interpretation and reproducibility. A potential way to fund these compliance checks is through a tiered, user-pays model of reproducibility. Under such a model, researchers who opt for higher levels of verification, such as submitting fully reproducible code and analyses for independent review, would pay more upfront but receive formal third-party verification of reproducibility, which could be recognised in funding and promotion decisions.

One of the main barriers to reproducing analyses was the difficulty in identifying which variables in the data corresponded to those used in the paper. Fewer than half of the studies with available data (20/43) included a data dictionary, and even when present, inconsistencies between variable names in the manuscript and those in the dictionary were common. This issue was particularly pronounced in studies with large numbers of variables, where identifying the correct inputs often involved a trial-and-error process, guessing likely variables and checking whether basic descriptive statistics (e.g., means or standard deviations) could be reproduced.

Datasets shared in statistical package formats such as SPSS or Stata were somewhat more likely to include inbuilt data dictionaries (e.g., variable labels and metadata), making it easier to match variables between the manuscript and the dataset. In contrast, datasets provided as unprocessed files (e.g., CSVs, Excel) often lacked this critical information unless a separate dictionary was supplied. Easily matching variables between the paper and the dataset is essential for reproducibility. Therefore, we recommend that authors provide data dictionaries that explicitly list the variable names used in the dataset, alongside the variable labels and descriptions as reported in the paper. In addition, we recommend that non-readable file formats, such as PDFs, Word and image files, should never be used to share data.

Reproducible code and data practices are essential for transparency, yet they remain uncommon in health and medical research [52]. Raising expectations of reproducibility must go hand in hand with formal training in reproducible workflows, not as an afterthought, but as a foundational skill set. This includes transparent coding, clearly structured data (e.g., long vs. wide format), and documented cleaning procedures. In our research, poor statistical reporting [24] and limited reproducibility were common, particularly when data cleaning steps were undocumented or unclear. These patterns may reflect a research culture shaped by publish-or-perish incentives, in which novelty and output volume are often rewarded over rigour and reproducibility [62].

Expectations can influence behaviour in ways that reinforce existing norms. The Pygmalion effect [63], which describes how individuals’ performance can be shaped by their expectations, offers a valuable lens for understanding how norms can become self-reinforcing. When transparency and documentation are not explicitly valued, and when it is assumed that health researchers lack the technical skills to implement reproducible practices, these low expectations can become self-fulfilling. Data sharing may become a tick-box exercise, and even shared data may lack the context required for meaningful reuse. Addressing this requires more than compliance with data sharing policies; it calls for cultural change supported by institutional incentives, clear journal requirements, and practical training. Embedding these expectations in early career researchers, particularly through academic programs and early career development, helps shift norms toward greater transparency and meaningful data reuse.

While the gold standard of reproducibility is for authors to provide code, it is recognised that many health researchers may not have the necessary training. To help improve reproducibility across all skill levels, we propose a Model Location and Specification Table (MLast) to accurately track analyses (Table 5). This table is metadata and could be presented in the supplementary material. For researchers with reproducibility skills, it could be automated by assigning statistical test labels in the same way tables or references are indexed, creating direct links between the paper, data and code. While code may improve reproducibility, it is often dense, requiring significant time and effort to track all the analyses. MLast provides a summary of analyses and would be a companion to any code. In the absence of code, the connection between the paper and the data is established by identifying the outcome variable in the paper and matching it to the variable names in the data using the model specification column. This connection will make the information machine-readable, thereby facilitating automated reproducibility.

**Table 5.**
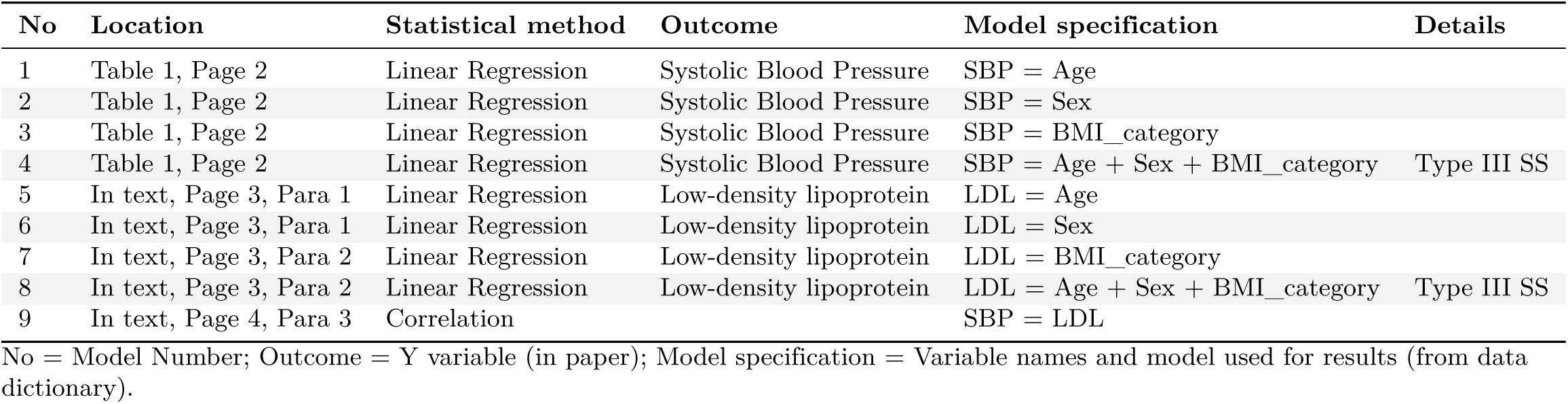
Hypothetical example of a model location and specification table.

For researchers without reproducibility skills, MLast can be completed manually and requires no additional skills beyond identifying the statistical tests used. Many researchers may find it challenging to identify all the statistical tests used in their paper, especially in health research, which can take several years. During that time, data and analyses may pass through multiple hands, making it essential to document the process. Clear links between the research questions and the reported results are also critical to ensure transparency and reproducibility and could be added to the details section of MLast or added as an extra column.

Greater automation may further improve computational reproducibility, with artificial intelligence (AI) potentially helping researchers develop, document, and retain statistical code during research [64]. However, these benefits depend on the quality of both the information provided to the AI and the statistical decisions underlying the generated analysis. AI-generated code may therefore improve documentation without necessarily ensuring that the analysis itself is appropriate or correct. The extent to which these tools perform reliably when used by researchers with varying levels of statistical expertise remains uncertain. Researchers with limited statistical expertise may also be less able to identify inappropriate statistical choices, errors in generated code, or analyses that do not adequately reflect the study design [65]. Early evidence suggests that AI-generated statistical analyses can be inconsistent and sometimes incorrect, with repeated analyses producing different results even for relatively straightforward statistical tasks [66]. Statistical literacy will therefore remain important as AI use in statistical analysis develops.

Applying AI retrospectively to reproduce published analyses presents additional challenges when the original analytical approach is poorly described, particularly when manuscripts report results without clearly identifying the statistical method used. Future research could directly evaluate how reliably AI can reproduce published statistical analyses, including whether it correctly identifies the intended analysis, makes appropriate statistical decisions, and reproduces the reported results.

### General recommendations for improving computational reproducibility

- Provide the dataset in a machine-readable format, such as Excel or CSV files.
- Provide data in a recognised format, either wide (where each participant has their own row) or long (where participants may have multiple rows).
- Create a reproducible analysis workflow and share the code used for data preparation, variable derivation, and statistical analysis.
- Specify the software environment used for the analyses, including the statistical software, version number, and key packages or libraries. This is often critical for computational reproducibility, particularly when defaults differ across versions.
- Provide a minimal, analysis-ready dataset containing the variables used in the statistical models, accompanied by clear documentation of any derived variables. Where complete analysis code is available, the source data and code used to create the analysis-ready dataset should also be shared.
- Provide a data dictionary that exactly matches the variable names used in the manuscript, including clear definitions, coding, and units where applicable.
- Ensure that the number of observations reported in the paper matches the dataset provided, and that the dataset corresponds to the final version used in the analyses.
- Clarify any data exclusions or pre-processing steps (e.g. filtering, recoding, transformations) applied before analysis, including the rationale and number of observations affected.
- Clearly describe the modelling process. The model type and key specifications (e.g. link function, random effects, transformations, clustering and selected variables) should be stated explicitly and summarised in table footnotes to improve transparency.
- Clearly describe how categorical variables were coded in models, including the reference category, and indicate this clearly in tables or footnotes. Variables with more than two categories should be represented using appropriate contrasts.
- Provide complete model results in the Supporting Information, including all analyses conducted, full regression outputs (coefficients, confidence intervals, p-values), and all variables included for adjustment.

### Comparison to other Reproducibility studies

Early efforts to assess computational reproducibility using shared data highlighted both the promise and the challenges of replicating scientific findings. For example, Ioannidis et al. [5] examined 18 microarray-based gene expression studies published in *Nature Genetics* (2005–2006), fully reproducing two and partially reproducing six. In their study, independent research teams assessed reproducibility, including aspects such as data processing, software details, and imaging parameters. Each team determined whether the main analysis results were (i) entirely reproduced in principle (all presented results were reproduced), (ii) partially reproduced (some results reproduced, others not), (iii) reproduced with discrepancies, (iv) partially reproduced with discrepancies, or (v) not reproduced at all. While the specific criteria and context differ, our study similarly emphasises graded reproducibility assessments rather than a binary reproduced/not reproduced assessment.

A study examining the reproducibility of code in machine learning research was conducted by Raff [67], who evaluated 255 papers published between 1984 and 2017. Reproducibility was defined as the ability to execute at least 75% of the original code to obtain the reported results, and 63% of the papers met this criterion. Only about half of the authors contacted responded, and successful reproduction was more common when authors engaged [67].

In contrast to Raff [67], we did not contact the authors in our study. Reproducibility was assessed solely based on the methods sections, results, and any supporting information provided with the papers. Although this could have included analysis code, none of the studies in our sample that provided data included code. This approach was intentionally adopted to reflect the experience of secondary users interpreting published findings independently. As such, our findings reflect the practical barriers faced by researchers attempting to reproduce results without author assistance, barriers that may be underestimated in studies that request additional information directly from the authors.

Laurinavichyute et al. [68] addressed the reproducibility challenge without direct author support. They recommended that authors verify whether their analyses can be reproduced solely from the materials they make available. Their study evaluated 59 papers published in the *Journal of Memory and Language* following the implementation of an open-data and code-sharing policy. They found 34% of papers were reproducible when accounting for rounding errors. This increased to 56% under more lenient criteria, allowing up to 20 deviations of up to 10%. The availability of analysis code was the strongest predictor of reproducibility, associated with a 40% higher probability of successful reproduction [68].

A study by Harris et al. [4] examined the reproducibility of descriptive statistics and odds ratios across six papers in public health services and systems research. Their study judged reproducibility based on the percentage difference for individual results, whether statistical significance changed, and differences in the mean width of confidence intervals. Paper-level differences were calculated as the mean and standard deviation of percentage differences. While each paper showed consistency in at least one of the four areas examined, inconsistencies were also common, primarily due to transcription errors and missing details about data management and analysis [4].

Although tolerance rules vary across computational reproducibility studies, prior frameworks have generally sought to distinguish trivial numerical variation from substantive discrepancies using predefined classification criteria. Hardwicke et al. [69] quantified discrepancies using relative error and classified them as minor or major based on percentage thresholds. Kambouris et al. [70] similarly used relative error but additionally introduced an explicit tolerance band derived from the unit of the last reported decimal place (10*−d*) to account for rounding precision. In a comparable manner, we derived two absolute tolerance thresholds from the number of decimal places (*d*) reported in the original paper (a stricter band: 0.5 *×* 10*−d*; a more lenient band: 0.999 *×* 10*−d*) and classified values as Reproduced, Incorrect Rounding, or Not Reproduced based on the absolute difference. This framework distinguishes plausible rounding variability from substantive numerical disagreement while remaining conceptually aligned with established computational reproducibility approaches.

### Limitations

A potential limitation of this study is that papers with more complete reporting may have had more opportunities to be classified as not reproduced compared with papers with minimal reporting. Although we adjusted for the probability of reproducibility classification using mixed multinomial regression to account for correlations within papers, this approach may not fully address the imbalance in reporting detail. Minor issues, such as typographical errors, were allowed within “Mostly Reproduced” models to account for limitations in reporting. Although incomplete reporting is closely linked to challenges in reproducibility, further work is needed to determine whether there is a systematic association between the extent of reporting and reproducibility outcomes.

The primary author assessed data sharing and computational reproducibility, with input from other authors and a bioinformatician as needed. This was an exploratory project, and the templates and methodology were refined during the initial stages of the study as challenges arose and were addressed. While some results may have been inadvertently missed, detailed record-keeping and a fully reproducible workflow in R Quarto helped minimise errors. Although only 20 papers were assessed for computational reproducibility, this sample size is consistent with many reproducibility studies, which are small because reproducing each paper typically requires one to three days. Importantly, the overall finding of poor reproducibility in health research was consistent with prior literature [5, 68], supporting the validity of the conclusions despite our refinement of the planned methods.

In some cases, poor reporting may have prevented the reproduction of results that could otherwise have been reproduced if the authors had been contacted. However, contacting authors was not undertaken, as the purpose of this study was to assess the extent to which results could be reproduced based solely on publicly available information and data. This approach reflects a broader goal: in the future, simple analyses are expected to become fully automated, based on accessible datasets and methods without the need for author involvement. Previous research has also shown that author follow-up can be time-consuming and often unsuccessful due to non-responsiveness [67]. This study presents a semi-automated framework in which original results required manual extraction, while the subsequent reproducibility checks were fully automated, providing a foundation for future advancements in this area.

### Conclusions

The reproducibility of health research is crucial for assessing the reliability of findings, identifying effective treatments, and improving patient care. Although data sharing is widely promoted to enhance transparency and reproducibility, these goals can be achieved only when shared data are complete, well documented, and usable. More than half of the studies assessed in this study were not computationally reproducible. Our study provides a framework for assessing computational reproducibility and demonstrates how these assessments can be implemented using reproducible templates. We also provide practical recommendations for authors, reviewers, and editors by specifying the materials and documentation required to enable independent computational reproduction of reported analyses.

Journals have an important role in improving computational reproducibility by enforcing data and code availability requirements rather than relying solely on author declarations. Editorial checks that analytical materials are available, accessible, and sufficiently documented could identify many reproducibility barriers before publication. More structured verification of these materials, as already implemented in some other disciplines, would likely further strengthen the reproducibility of published health research.

Where published results cannot be computationally reproduced, their reliability remains uncertain. Further investigation may show that a discrepancy can be resolved by clarifying undocumented analytical details, or may identify an error that materially affects or invalidates the original conclusion. Such results should not be relied upon for clinical or medical decision-making until the underlying discrepancy has been resolved, and should be corrected where errors are confirmed. Computational reproducibility alone does not establish that an analysis is statistically appropriate or correctly interpreted; these issues require separate evaluation of the statistical approach and its assumptions.

We recommend that researchers share the code used for their analyses, allowing reported results to be directly reproduced from shared data and, where possible, generated automatically. While full reproducibility remains a longer-term goal and many researchers may not yet have the coding skills required, we propose a Model Location and Specification table (MLast) as a practical mechanism for improving transparency. MLast records where analyses were performed and how models were specified and can be created by researchers across a range of skill levels, including through automated approaches. When used alongside a data dictionary, it provides metadata linking analyses to the corresponding data and gives a clear overview of what was modelled and how. MLast may also provide an important foundation for future automated reproducibility assessments by enabling data and reported analyses to be more readily matched.

Eliminating all errors from published research is unlikely to be achievable. Reproducible research should instead aim to minimise errors and ensure that they can be identified and corrected transparently through peer review or post-publication review. Recognising that scientific papers are not necessarily static, some have advocated treating them as “living” documents that can be updated to address reproducibility problems or correct errors [67]. This perspective reflects the dynamic nature of science and emphasises shared responsibility for maintaining the accessibility, reproducibility, and reliability of research outputs. Achieving this will require greater investment in researcher training in reproducible practices and continued support for meta-research to develop and evaluate strategies for improving statistical and research quality. Responsibility for reproducibility should therefore not rest solely with individual authors or journals, but should be supported by research institutions, funding bodies, and publishers.

## Data Availability

The data and a reproducible R Quarto file used to produce this paper, including tables, figures, and code, have been stored in a GitHub repository and can be cited using Zenodo dio.

https://github.com/Lee-V-Jones/Reproducibility

https://doi.org/10.5281/zenodo.19448969

## Acknowledgements

We acknowledge Adrian llich for providing bioinformatics We thank all the statisticians who generously gave their time to review papers for this study, including those acknowledged by name below and others who chose to remain anonymous: including: Ingrid Aulike, Peter Baker, Brigid Betz-Stablein, Enrique Bustamante, Taya Collyer, Susanna Cramb, Alanah Cronin, Laura Delaney, Zoe Dettrick, Eralda Gjika Dhamo, Des FitzGerald, Peter Geelan-Small, Edward Gosden, Alison Griffin, Jenine Harris, Cameron Hurst, Kyle James, Helen Johnson, Jessica Kasza, Karen Lamb, Stacey Llewellyn, James Martin, Miranda Mortlock, Satomi Okano, Alan Rigby, Michael Steele, Megan Steele, Jacqueline Thompson, Simon Turner, Michael Waller, Kevin Wang, Jace Warren, Natasha Weaver, Lachlan Webb, and Janet Williams.

## Transparency and Declarations

Version 1 of this paper was peer reviewed by MetaROR (DOI: 10.70744/MetaROR.417.1.rv1). The current Version 2 incorporates revisions made in response to that peer review.

## Author contribution statement

- Lee Jones: Study design, analysis, software, writing original draft, manuscript review and editing.
- Adrian Barnett: Study design, contribution to results validation, manuscript review and editing.
- Gunter Hartel: Contribution to results validation, manuscript review and editing.
- Dimitrios Vagenas: Study design, contribution to results validation, manuscript review and editing.

## Competing Interest Statement

The authors have declared no competing interest.

## Funding Statement

There was no cost associated with this research except for attending conferences. These costs were covered by the primary authors PhD allocation from the health faculty, Queensland University of Technology, and scholarships. The Statistical Society of Australia (SSA) and the Association for Interdisciplinary Meta-research & Open Science (AIMOS) supported the primary author with travel grants to attend their respective conferences. These scholarships did not influence the results of the study. The funders had no role in study design, data collection and analysis, decision to publish, or preparation of the manuscript.

## Supporting Information

**Table S1.** Reproducibility of reported model statistics.

| Terms | Reproduced | Incorrect Rounding | Not Reproduced |
| --- | --- | --- | --- |
| B | 0.51 (0.45, 0.57) | 0.06 (0.03, 0.11) | 0.43 (0.37, 0.48) |
| Lower CI | 0.60 (0.51, 0.68) | 0.10 (0.04, 0.17) | 0.31 (0.24, 0.38) |
| Upper CI | 0.57 (0.49, 0.66) | 0.07 (0.03, 0.13) | 0.36 (0.28, 0.43) |
| SE |  |  |  |
| t |  |  |  |
| p-value | 0.54 (0.47, 0.60) | 0.04 (0.02, 0.08) | 0.42 (0.37, 0.48) |
| $R^2$ | | | |
| Adjusted $R^2$ | | | |
| F |  |  |  |
| Standardized B |  |  |  |

Terms = model statistics; Predicted probabilities with 95% credible intervals were estimated using a Bayesian multinomial mixed-effects model with a random intercept for paper. Models were estimated for the terms with sufficient data. Predicted probabilities were not calculated for terms with $N < 30$ .

**Table S2.** Observed frequencies and predicted probabilities for reproducibility of models.

| Category | n (%) | Predicted probability |
| --- | --- | --- |
| Reproduced | 12 (24%) | 0.23 (0.03, 0.59) |
| Mostly reproduced | 15 (29%) | 0.30 (0.08, 0.64) |
| Partially reproduced | 5 (10%) | 0.10 (0.01, 0.35) |
| Not reproduced | 19 (37%) | 0.37 (0.09, 0.73) |

Category = The reproducibility status for models; n (%) = observed frequencies and percentages of reproduced models. Predicted probabilities = The predicted probabilities with 95% credible intervals estimated using a Bayesian multinomial mixed-effects model with a random intercept for paper.

## Notes

### Author Declarations

The aim of this study was to reproduce the statistical results from publications that made their data publicly available. Authors of papers published in PLOS ONE were considered to have implicitly consented through their agreement with the journals data sharing policy, which supports the validation and reproduction of results from shared data. This study received Negligible-Low Risk Ethics approval from the Queensland University of Technology Human Research Ethics Committee (Approval Number: 2000000458).

### Summary of Updates

This version of the manuscript has been revised in response to the review from MetaROR.

